# Outcomes associated with SARS-CoV-2 viral clades in COVID-19

**DOI:** 10.1101/2020.09.24.20201228

**Authors:** Kenji Nakamichi, Jolie Z. Shen, Cecilia S. Lee, Aaron Lee, Emma A. Roberts, Paul D. Simonson, Pavitra Roychoudhury, Jessica Andriesen, April K. Randhawa, Patrick C. Mathias, Alex L. Greninger, Keith R. Jerome, Russell N. Van Gelder

## Abstract

**Background:** The COVID-19 epidemic of 2019-20 is due to the novel coronavirus SARS-CoV-2. Following first case description in December, 2019 this virus has infected over 10 million individuals and resulted in at least 500,000 deaths world-wide. The virus is undergoing rapid mutation, with two major clades of sequence variants emerging. This study sought to determine whether SARS-CoV-2 sequence variants are associated with differing outcomes among COVID-19 patients in a single medical system.

**Methods:** Whole genome SARS-CoV-2 RNA sequence was obtained from isolates collected from patients registered in the University of Washington Medicine health system between March 1 and April 15, 2020. Demographic and baseline medical data along with outcomes of hospitalization and death were collected. Statistical and machine learning models were applied to determine if viral genetic variants were associated with specific outcomes of hospitalization or death.

**Findings:** Full length SARS-CoV-2 sequence was obtained 190 subjects with clinical outcome data. 35 (18.4%) were hospitalized and 14 (7.4%) died from complications of infection. A total of 289 single nucleotide variants were identified. Clustering methods demonstrated two major viral clades, which could be readily distinguished by 12 polymorphisms in 5 genes. A trend toward higher rates of hospitalization of patients with Clade 2 was observed (p=0.06). Machine learning models utilizing patient demographics and co-morbidities achieved area-under-the-curve (AUC) values of 0.93 for predicting hospitalization. Addition of viral clade or sequence information did not significantly improve models for outcome prediction.

**Conclusion:** SARS-CoV-2 shows substantial sequence diversity in a community-based sample. Two dominant clades of virus are in circulation. Among patients sufficiently ill to warrant testing for virus, no significant difference in outcomes of hospitalization or death could be discerned between clades in this sample. Major risk factors for hospitalization and death for either major clade of virus include patient age and comorbid conditions.

**Funding:** Supported by NIH P30EY001730, the Mark J. Daily, MD Research Fund (RVG), the Alida and Christopher Latham Research Fund (RVG, AYL, CSL), NIH K23EY029246 (AYL), US Food and Drug Administration (QYL)

## Introduction

Coronaviruses are a group of enveloped, non-segmented, single-stranded, positive-sense RNA viruses that are capable of infection in humans and animals ^1^. SARS-CoV and MERS-CoV are two coronaviruses that have previously resulted in large-scale pandemics ^2^. COVID-19, the disease caused by the coronavirus SARS-CoV-2, has affected over 10 million people and resulted in over 500,000 deaths worldwide in a seven-month period beginning in December 2019 (https://coronavirus.jhu.edu/map.html, accessed 7/31/20).

Clinical outcomes of COVID-19 vary significantly. Overall, worldwide mortality is approximately 6% of clinically confirmed cases, while hospitalization rates in the US average approximately 15% of such cases (https://coronavirus.jhu.edu/map.html) (although given the high prevalence of asymptomatic or unreported infections, these values likely overestimate mortality and hospitalization rate substantially ^3^). While age and comorbid conditions have been identified as risk factors for hospitalization and death ^4-6^, outcomes appear variable even among large populations. For example, at present, comparing outcomes from two large states in the US (New York and California), the former has a nearly 8% mortality rate among confirmed cases compared with a 3.4% rate in the latter (https://coronavirus.jhu.edu/map.html). Rates of hospitalization also differ substantially between regions.

SARS-CoV-2 is ∼30kb in length and contains 16 open reading frames (ORFs) ^7,8^. Despite its very recent emergence as a viral pathogen, SARS-CoV-2 has undergone rapid mutation. The Nextstrain sequencing consortium ^9^ has identified 2785 variants, while the GISAID initiative has collected over 50,000 full length sequences as of 6-28-20 ^10-12^. Analysis of these sequences reveals 5 (Nextstrain) or 6 (GISAID) sequence clusters, with as many as 18 new mutations in each cluster distinct from the original SARS-CoV-2 virus.[6]

Viral genetic variation plays an important role in pathogenicity and virulence in other viruses, such as influenza ^13^. In this study, we sought to determine if specific viral sequence variants are associated with better or worse clinical outcomes in COVID-19, by analyzing the clinical course of a cohort of patients within a single medical system for whom both full-length viral sequence data and clinical outcome data were available.

## Results

### Study population characteristics

Full length SARS-CoV-2 sequence was obtained from 283 patients; clinical history within the UW Medicine system was available from 190 of these. 35 (18.4%) were hospitalized and 14 (7.4%) died from complications of the infection. Clinical characteristics of this cohort are summarized in Table I. The mean age of all COVID-19 patients, regardless of hospitalization status, was 53.4 (range 16-95), 51.1% of patients were male, and 14.8% came from a skilled nursing facility (SNF). Of all patients, 56.3% were white, 11.6% Black, 11.1% Asian, 1.6% American Indian/Alaskan Native, 1.1% Native Hawaiian/Pacific Islander and 18.2% other race or ethnicity. The most common comorbidities seen in our patient cohort were hypertension (34.1%), history of cancer (18.4%), cardiovascular disease (CVD) (18.4%), diabetes (16.8%), asthma (12.4%), and hypothyroidism (12.4%). Less prevalent were chronic kidney disease (CKD) (8.1%), chronic heart failure (CHF) (6.5%), chronic obstructive pulmonary disease (COPD) (4.9%), a history of deep venous thrombosis (DVT) (4.3%), and previous myocardial infarction (MI) (3.2%). With regard to medication use, 15.8% of all COVID patients were taking corticosteroids or immunomodulatory therapy, 20.1% were taking an angiotensin converting enzyme (ACE) inhibitor or angiotensin receptor blocker (ARB), and 10.9% were on chronic anticoagulation. 10.0% of patients were current smokers.

When stratified between hospitalized and non-hospitalized patients, advanced age, admission from a skilled nursing facility, and a history of either HTN, CHF, CVD, CKD, and a history of DVT or cancer were significantly associated with hospitalization (p-values <0.001 - 0.02). Other characteristics that were associated with hospitalization included anti-coagulated status, utilization of an ACE inhibitor or ARB, and use of corticosteroid or immunomodulatory therapy (p-values <0.001 - 0.04). Notably, comorbidities of diabetes and known tobacco history was not significantly associated with hospitalized status (Table 1).

**Table 1.**
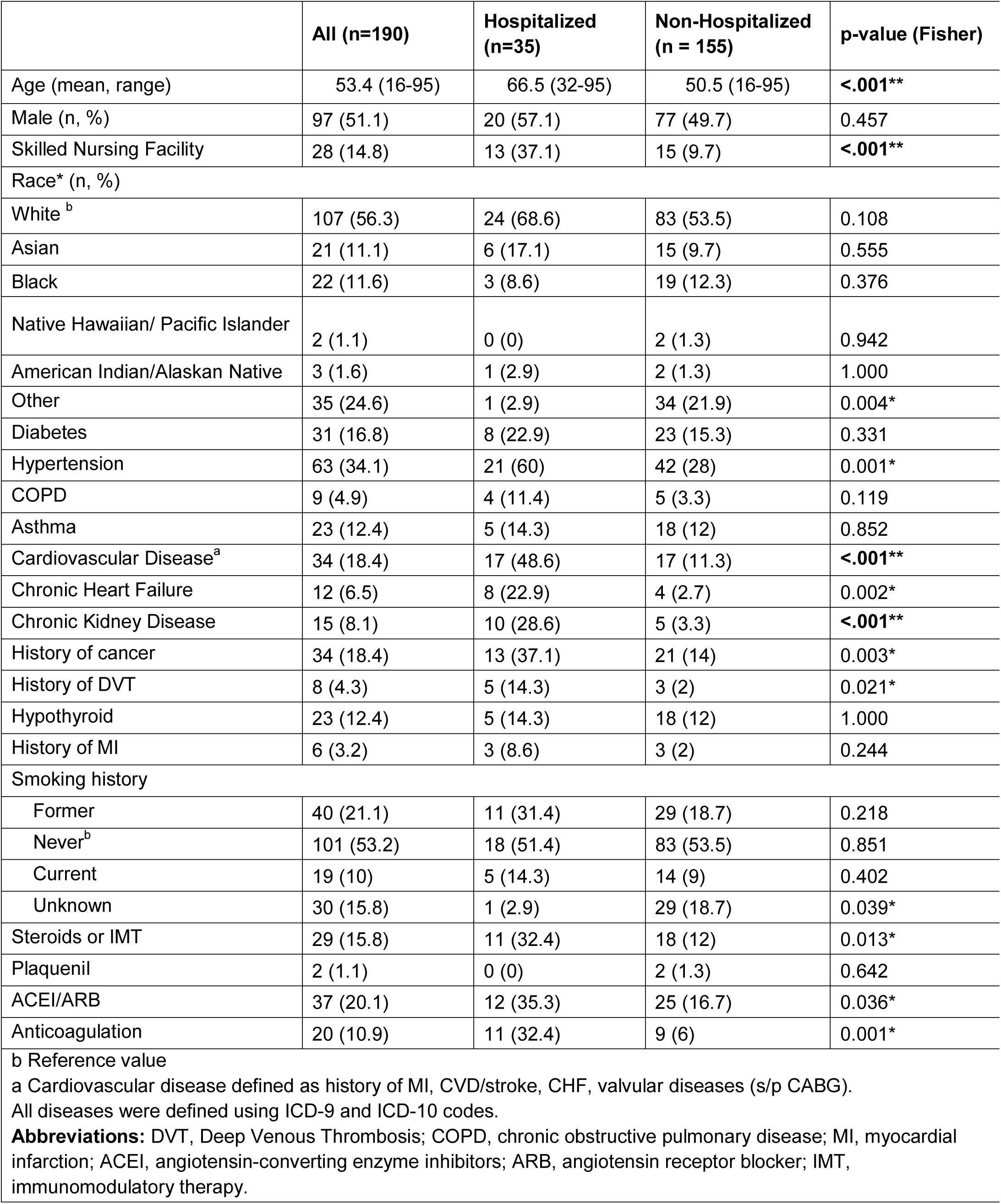
Baseline demographics and medical histories of hospitalized and non-hospitalized patients

### Viral sequence variants

Full-length sequence was obtained from 190 samples acquired from UW Medicine sites in Seattle Washington between 3/1/20 and 4/15/20. Relative to reference SARS-COV-2 sequence, these samples in aggregate showed 289 sequence variants (283 SNPs, 6 insertion/deletion, Figure 1). Most variants were present with frequency less than 5%. 163 of the sequence variants were missense mutations, 84 were synonymous mutations, and the remainder were not in protein coding regions.

**Figure 1.**
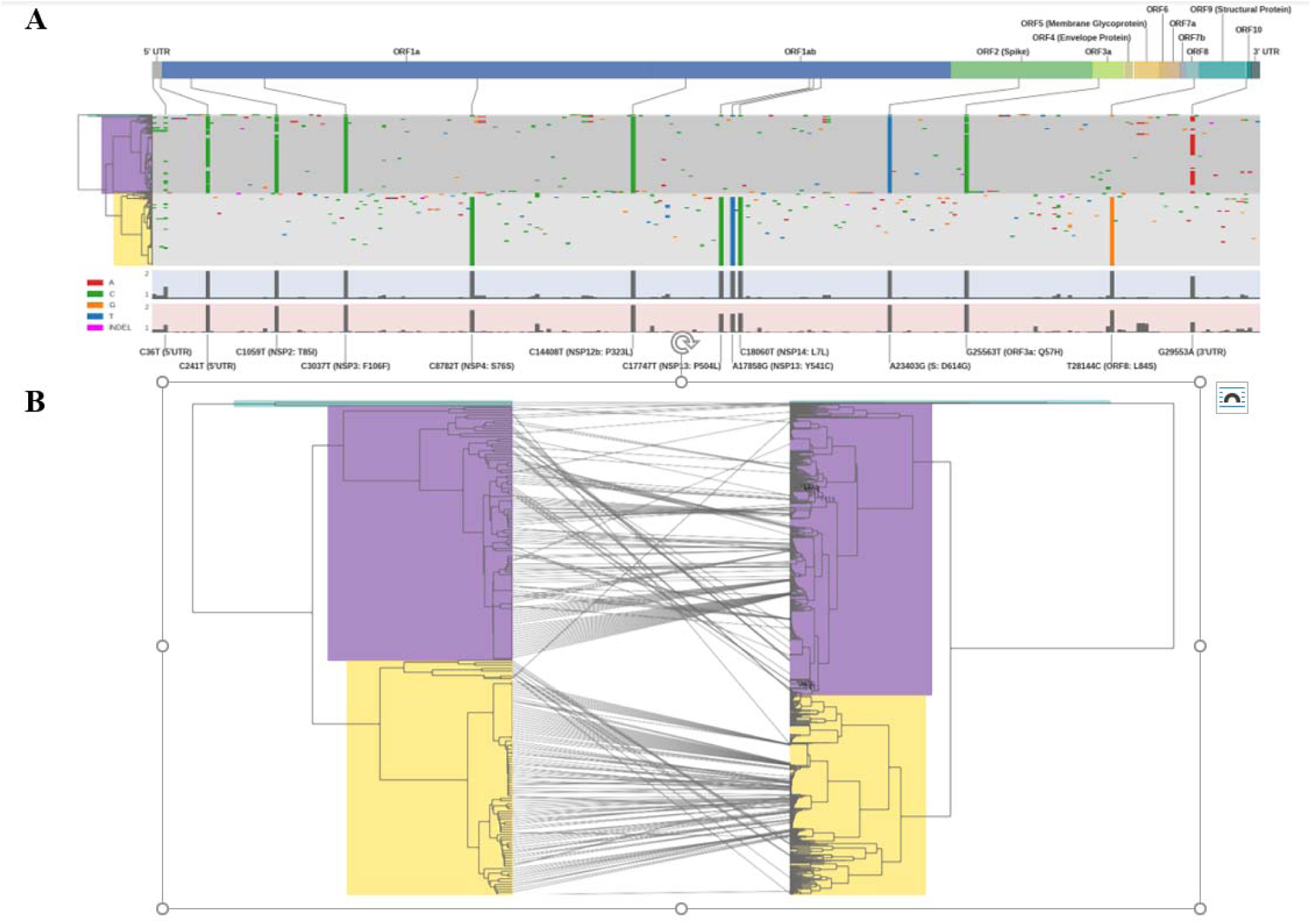
Top – SARS-CoV-2 sequence variants among 190 full length genomes sequenced from outbreak in Seattle, WA March-April 2020 Purple = Clade 1, Gold = Clade 2. Bottom: dendrogram of sequence relations, mapped to identical analysis of 2753 full length sequences in NCBI database.

UPGMA hierarchical clustering produced two clear clades of sequence variants (Figure 1), determined by 12 single nucleotide polymorphisms (Table 2). Ninety-seven samples corresponded to what we refer to as ‘Clade 1’ and 91 corresponded to ‘Clade 2’. Two of 190 samples did not fall into either of the two major clades. When mapped onto GISAID and NextStrain clades, we find in clade 1 that 89 correspond to clades GH/20C, 6 map to G/20A, and 2 map to G/20B. In clade 2 we that that 86 correspond to S/19B, and 5 mapped onto L/19A (Table 3). The 2 of 190 samples that did not fall into either of the major clades corresponded to GH/20C and S/19B.

**Table 2.**
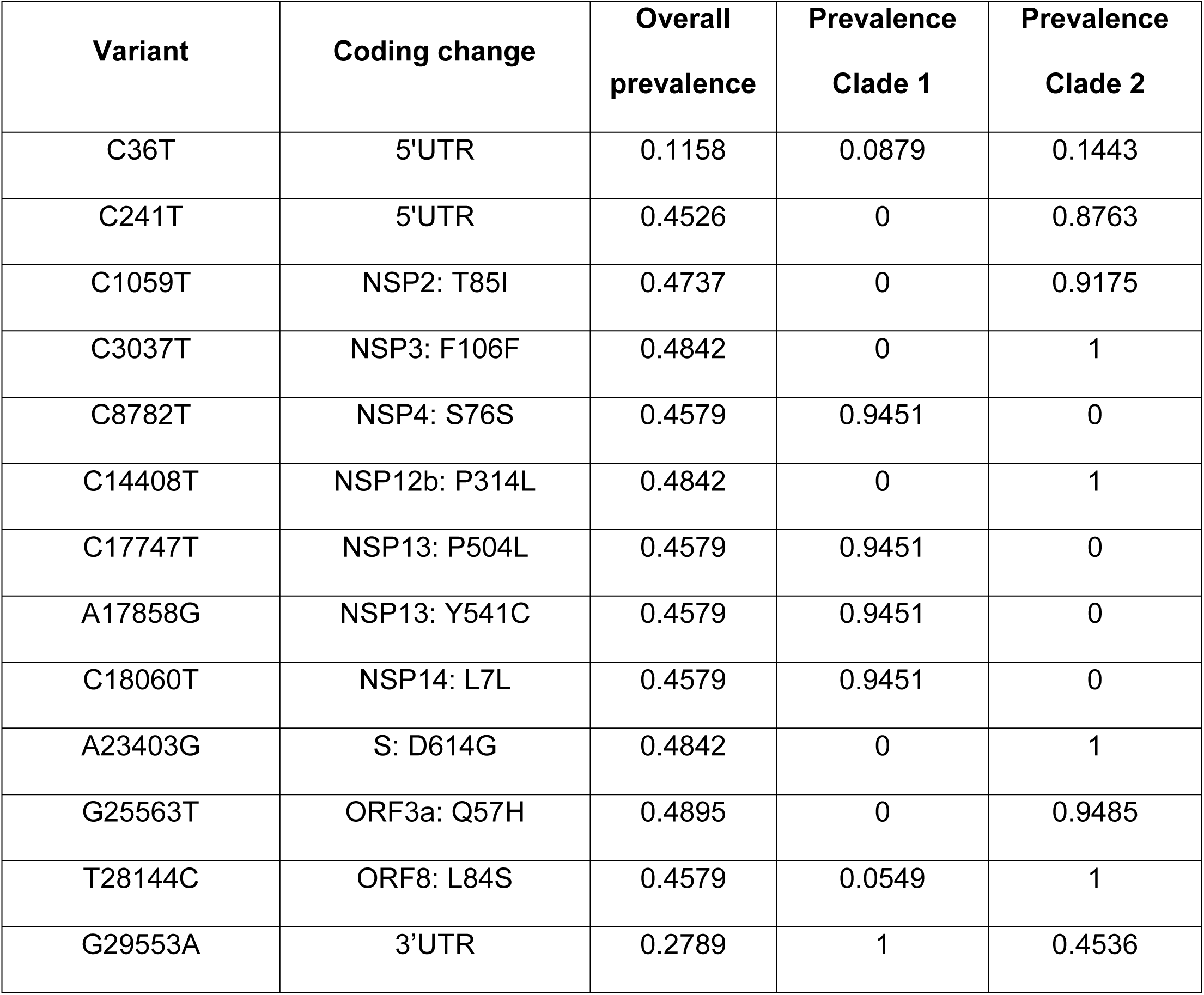
Distribution of sequence variants occurring more frequently than 5% in the study population.

**Table 3.**
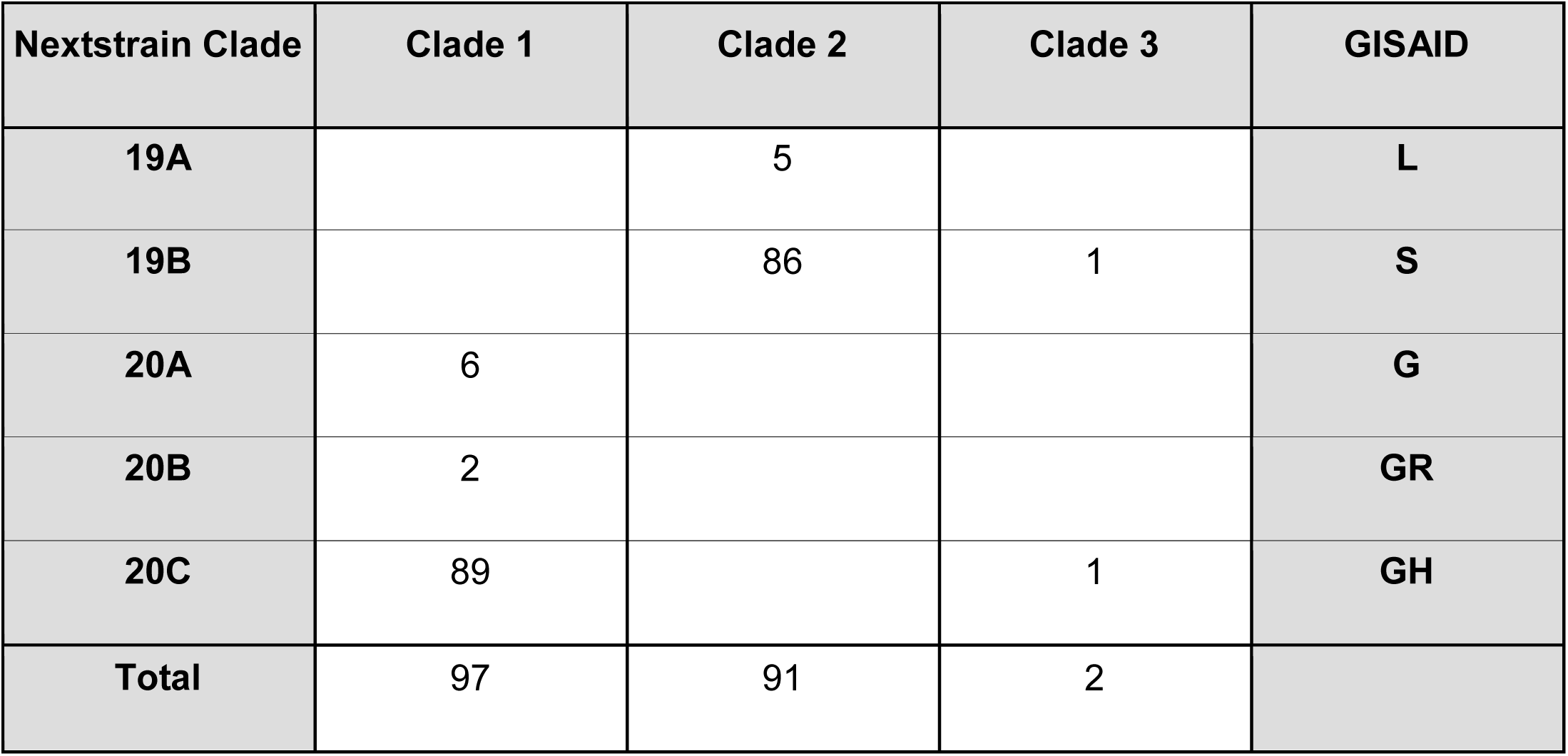
Correspondence of Clade 1 and Clade 2 of current cohort to Nextrain and GISAID clades

Mapping the 190 sequence variants onto the 2563 available full-length sequences in NCBI Virus (05/18/2020) showed that the sequence variants found in Seattle in this study represented a substantial fraction of sequence variation seen globally (Figure 1B). Of note, the two major clades identified in our Seattle-based samples are found in approximately equal proportion among global samples. No variants were observed in the current series that did not map on an existing clade. In Seattle, there was underrepresentation of one clade appearing in the larger dataset, which included sequences collected from 20 countries across 5 continents including the USA, China, Australia, Italy, Pakistan, and Brazil and contained the reference sequence (NC_045512.2). Overall, however, the sequences variants obtained in this study did not appear unique to Seattle, and represented a substantial proportion of variation noted globally.

To determine if differential selective pressure was driving mutagenesis toward one or the other major clade, the number of missense to silent mutations was compared between clades. Overall, the ratio of missense to sense was 1.851 in Clade 1 and 1.889 in Clade 2. In each case, this ratio varied significantly from expected random mutation which would have predicted a ratio of 3.46 (p < 0.001 for each). Thus, both clades appear to be under selective pressure but neither appears under differential pressure.

### Correlation of sequence variants with clinical outcomes

Viral clade appeared to correlate with several baseline clinical characteristics as shown in Table 4. When stratified between clade 1 and clade 2 infections, there was a significant difference in patients with a history of CVD, malignancy, steroid/IMT use, and anti-coagulation as well as in patients with a smoking history (p-values: 0.005 – 0.03).

**Table 4.**
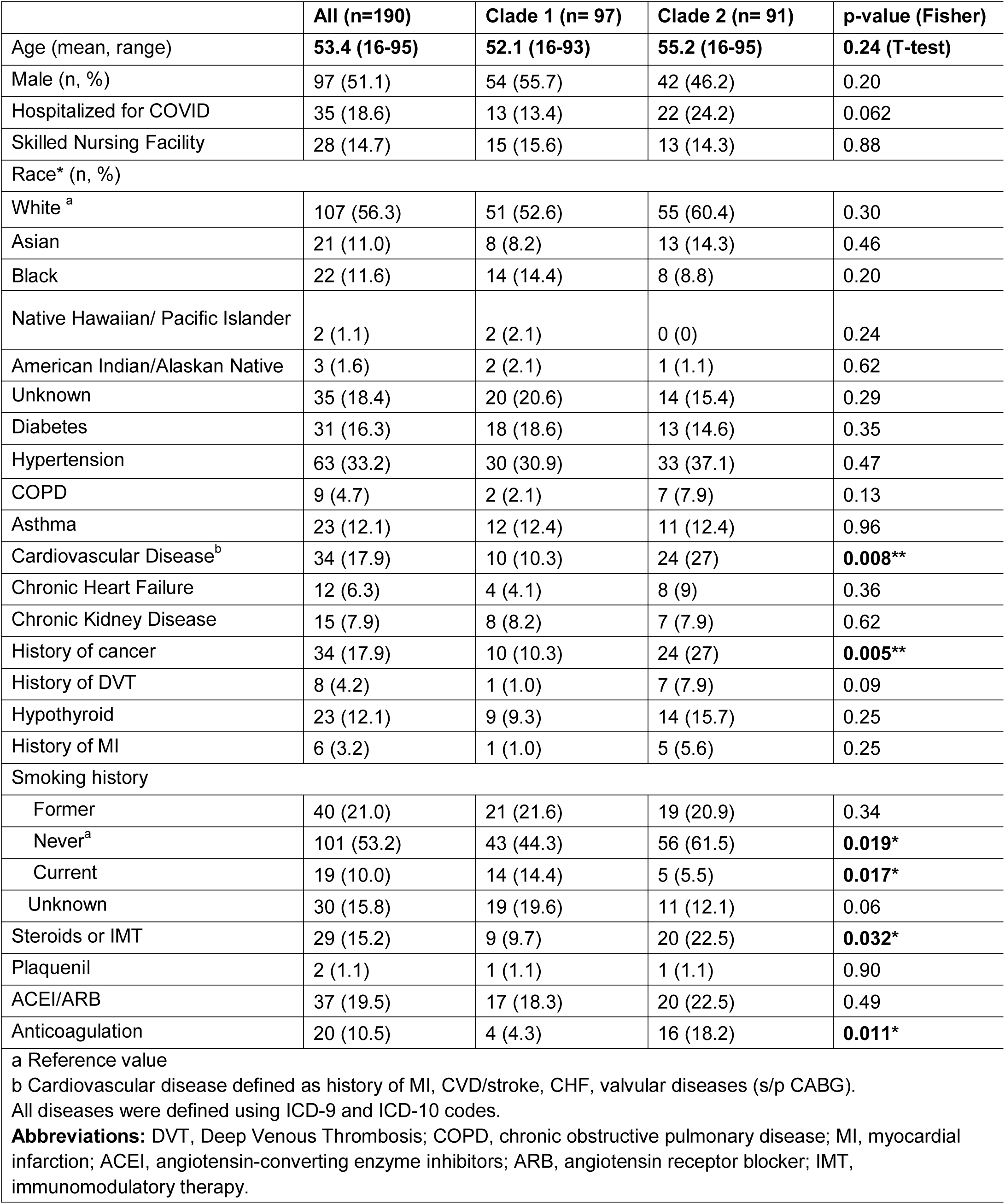
Demographic Factors and Baseline Clinical Characteristics of the Study Population Stratified by Viral Clade (n=188)^a^.

Comorbidity with CVD and cancer history was associated with clade 2 infection (OR: 3.1, 3.1, respectively). History of steroid/IMT and anticoagulation use was associated with clade 2 infection (OR: 2.7, 5.0 respectively). Notably, patients who had never smoked tobacco were more likely to be infected with clade 2 (OR: 2.0) while patients who were active smokers were more likely to be infected with clade 1 (OR: 3.6). In multivariable analyses, a history of malignancy was significantly associated with clade 2 across all five feature selection models (aOR: 3.4; p-values: .009 - .04) A current smoking history was significant associated with clade 1 in all multivariate models except for the all-variable model (aOR: 4.1, p-values: 0.01 – 0.07)

Outcomes of patients by viral clade are shown in Figure 2. 13/97 (13.4%) patients from Clade 1 were hospitalized compared with 22/91 (24.2%) of Clade 2 (p = 0.063 by Fisher exact). 6/97 (6.5%) patients from Clade 1 died from infection compared with 8/91 (8.8%) from Clade 2 (p = 0.58 by Fisher exact). Further analysis of individual viral sequence polymorphism revealed that no single polymorphism was significantly associated with outcomes of hospitalization or death, even without statistical adjustment for multiple comparisons.

**Figure 2.**
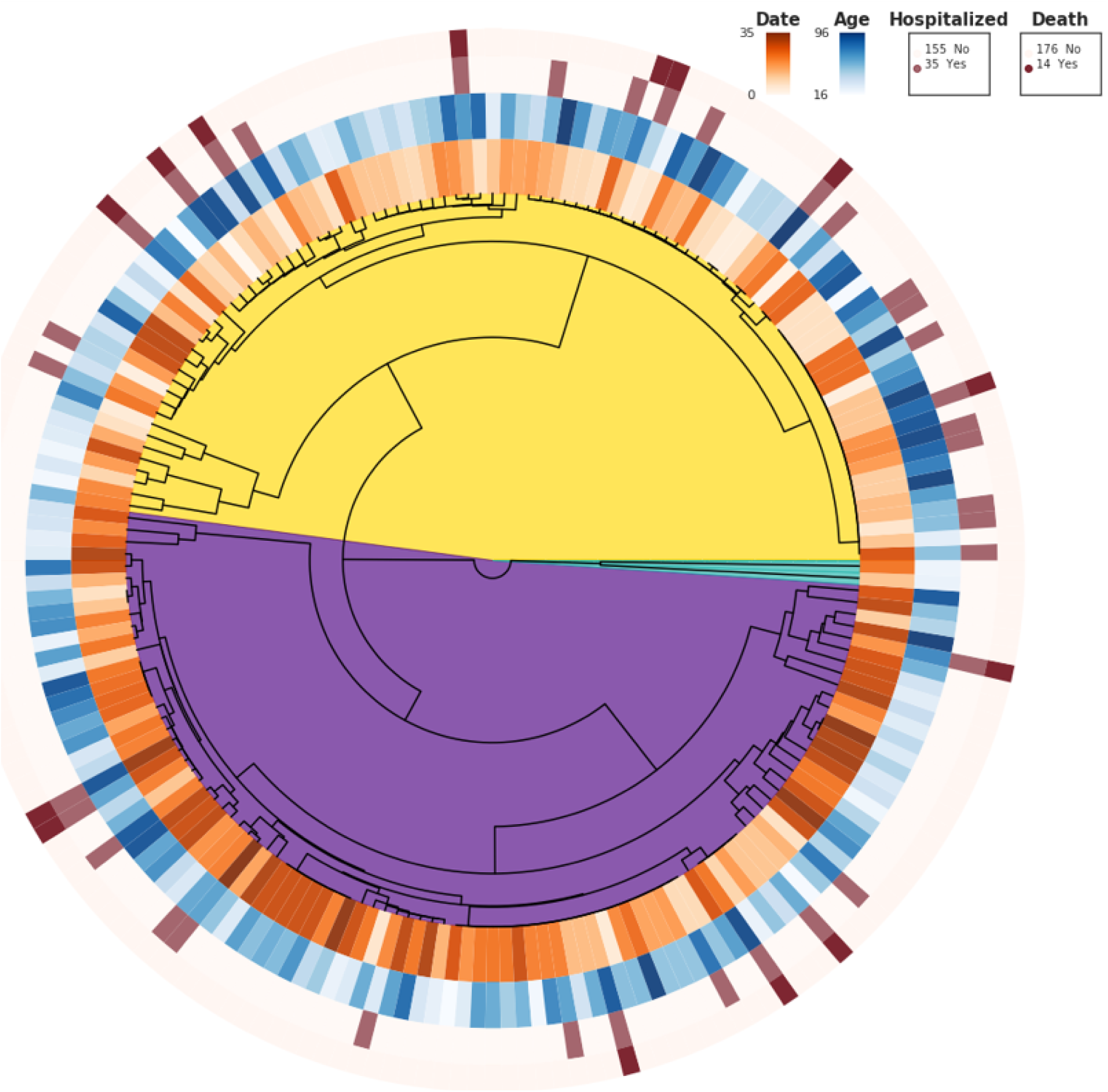
Outcomes of COVID-19 in cohort, divided by viral clade. Color code and dendrogram as in Figure 1. Date refers to relative date of sample acquisition over 35 days, darker color is more recent.

Machine learning (ML) was applied to probe for cooperative effects between viral genotype and host risk factors in determining which patients would be hospitalized. Death was too infrequent an outcome to allow machine learning modeling from our dataset. Using a Random Forest (SKLearn ^14^) we trained models using patient demographics, clinical features, and viral clade either separately or in combination on 160 cases, and tested the model on a hold-out set of 30 cases. For our initial analysis, we utilized the most recent 30 cases (which had 3 hospitalized patients out of 30). As shown in Figure 3, the model using solely patient demographics achieved an AUROC of 0.66. Addition of clinical information to demographics resulted in a substantially improved AUROC of 0.93. This model correctly predicted hospitalization status in 26 of 30 patients. Addition of clade data to either demographics-only or demographics+clinical models resulted in minimal improvement (AUROC 0.72 and 0.93, respectively). A final model, in which all genetic polymorphism data was added to the demographics+clinical+clade model performed no better than the demographics+clinical model (AUROC 0.89).

**Figure 3.**
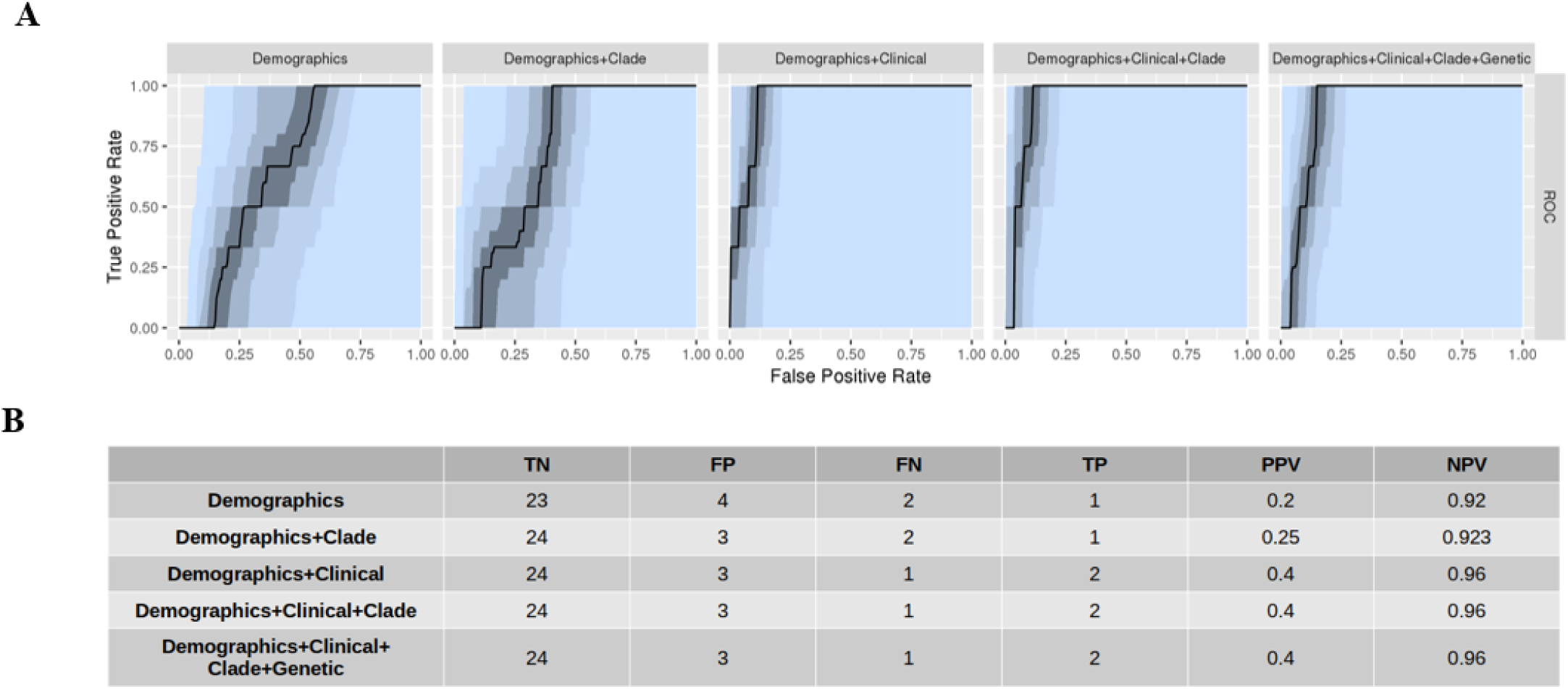
AUROC for machine learning models for prediction of hospitalization using test set of 30 most recent cases. Bottom: Optimal model performance for each dataset model for prediction of hospitalization.

Because hospitalization was relatively rare (18% in total data set), machine learning models could succeed by generally predicting non-hospitalization. The best trained model in our initial analysis (demographic + clinical) correctly predicted hospitalization status in 26 of the 30 subjects. The negative predictive value of this model was excellent (0.96), but the pre-test probability of non-hospitalization in this cohort was already 0.9. To test the generalizability of the machine learning approach for a group with higher risk of hospitalization, we re-trained the model on the same data, except holding out the most recently collected 15 hospitalized and 15 non-hospitalized patients (Figure 4). Again, AUROC of the demographics-only model was relatively high (0.78). Performance was again improved with addition of clinical data (AUROC 0.86), and no further improvement was seen with addition of clade or genetic data to the demographics+clinical dataset. Interestingly, with the 50% hospitalization holdout set, a demographics + clade model did appear to out-perform demographics-only (AUROC 0.86), suggesting that there may be some outcome information associated with clade, with the model correctly predicting two additional hospitalizations and one non-hospitalization. However, addition of clade information to the demographics + clinical did not improve performance further, suggesting minimal interaction between viral clade and comorbid conditions.

**Figure 4.**
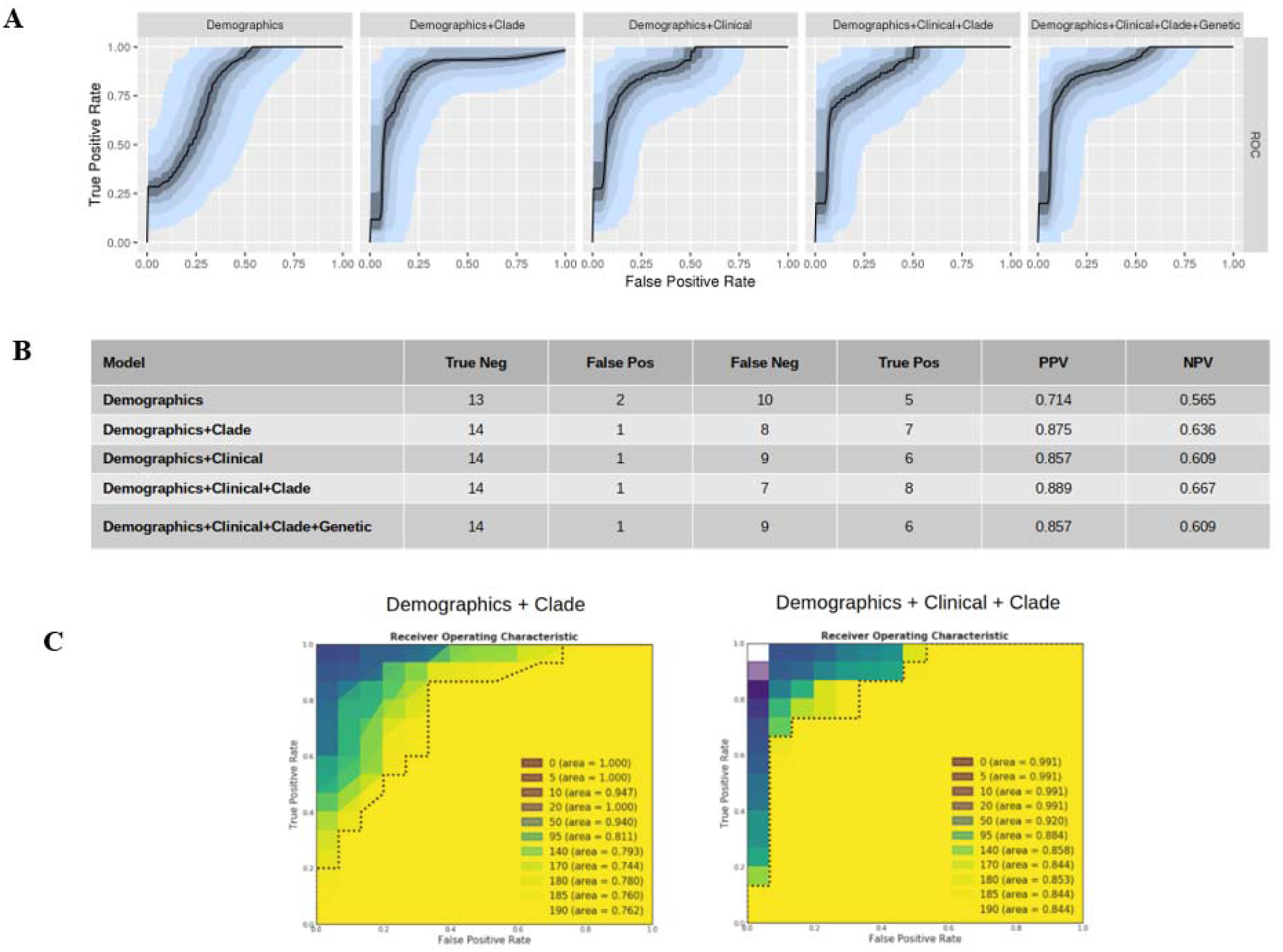
Machine learning models for prediction of hospitalization using hold-out set of 15 most-recent hospitalizations and 15 most-recent non-hospitalizations. A: AUROC for machine learning models on hold-out set. B: Optimal model performance for each dataset model for prediction of hospitalization. C: Model AUROC for spiked dataset ranging from 100% strain concordance with hospitalization (0) to complete randomization of outcome with respect to clade (190). Note that models approach observed AUROC with 50/160 randomized with respect to clade.

We further tested the machine learning models for sensitivity by training the models on datasets in which a specific clade had a ‘spiked’ association with hospitalization, ranging from 100% to random assignment (Figure 4C). Spiked models achieved AUROC of 1.0 for clade assignments ranging from perfect correlation to ∼10% noise, but then degraded toward demographic-only level AUC performance with between 25% and 50% noise. This demonstrates that had viral clade been a dominant determinant of outcome the machine learning models would have had sufficient power to detect this effect.

## Discussion

The COVID-19 pandemic of 2019-2020 has had a dramatic impact on health world-wide, with 7.8 million cases and over 400,000 attributed deaths world-wide as of June 14, 2020. The first case reported in the United States was in the state of Washington in January 2020. In the ensuing 6 months, over 20,000 cases have been reported in Washington. The overall hospitalization rate within Washington State as of 7/31/20 is approximately 15%, with an overall mortality rate of 5% (https://coronavirus.jhu.edu/map.html), although these numbers are based on Department of Health confirmed cases and likely overestimate true population rates (which would include asymptomatic and minimally symptomatic infected individuals).

Several possibilities exist for the heterogeneity of outcomes associated with SARS-CoV-2 infection. Morbidity may be associated with pre-existing illness; individuals already ill from other causes may be more likely to require hospitalization or succumb from infection. Access to quality healthcare may also influence outcomes. Age itself is a predictor of hospitalization and mortality which may significantly influence outcome. Host genetic factors are likely to play a role as well, and recent results suggest two host susceptibility loci that influence outcome in COVID-19 ^15^.

For some infectious processes, viral strain may play a large role in determination of pathogenicity. The influenza pandemic of 1918 – the last widespread highly morbid global pandemic – was caused by a specific strain of influenza virus (H1N1). Many other viruses have significant sequence variation which influences clinical outcomes ^16-18^. Hepatitis C genomic variants, for example, have been significantly associated with outcomes in hepatic disease ^19^.

The current study is the first to attempt to link SARS-CoV-2 viral sequence variants with COVID-19 outcomes. We found significant variation in viral sequence in our sample, with 289 sequence variants found in the 190 sequenced samples. However, the majority of variants occurred with a frequency of less than 10%. All of the frequently encountered variants have been previously described, and several clades have been identified in the literature ^20-24^.

Within our data, two clear clades emerged from UPGMA hierarchical clustering of the sequence variants. The clades are determined by 12 base variants as noted in Table 2. 97 samples corresponded to what we refer to as ‘Clade 1’ and 91 corresponded to ‘Clade 2’. When mapped onto GISAID and Nextstrain clades, we find in clade 1 that 90 correspond to clades GH/20C (GISAID/NextStrain), while in clade 2, 86 correspond to S/19B. In analyzing the ratio of synonymous to non-synonymous mutations, we find that both major clades appear to be under substantial negative selection, with significantly more synonymous than non-synonymous mutations observed than would be observed by chance. However, relative to each other, neither strain appeared to be under differential selective pressure.

We find that risk factors for hospitalization for patients with COVID-19 include advanced age and presentation from skilled nursing facility. In addition, we found that histories of hypertension, cardiovascular disease, deep venous thrombosis, and chronic renal disease were associated with hospitalization. Even though we found several baseline clinical factors to be significantly associated with clade 2 in univariate analyses and history of malignancy in the multivariate model, rates of hospitalization were not significantly different between patients infected with the two major clades of virus in our study (p=0.063), nor were mortality rates (p=0.58). Given the relatively low number of fatalities in our study, we were not powered to detect subtle strain-level differences in mortality outcome.

Machine learning approaches allowed us to model the predictability of hospitalization. Demographics alone was sufficient to allow some prediction of hospitalization with an AUROC for the model for the most recent 30 cases of 0.66. However, addition of clinical data improved the AUROC to 0.93. Addition of clade or individual viral sequence data to the model did not further improve performance suggesting that viral sequence variants do not independently contribute significantly to risk of hospitalization. Sensitivity analysis suggests that had a viral variant had >50% impact on hospitalization risk this would have been detected by the machine learning algorithm and resulted in higher AUROC.

This study has some unique strengths and weaknesses. Our data were derived from a single health-care system encompassing three hospitals in a major metropolitan area. By using a single medical system, we had access to substantial medical history on these subjects as well as reduced concern regarding the influence of hospital system on outcomes (i.e. we assume that decisions for hospitalization and quality of care of hospitalized patients will be more consistent in patients treated within a single system). Our system served as the primary site for COVID-19 testing particularly in March and April, 2020, which gave us access to a substantial number of patients with linked outcome data. However, our outcomes at present are limited to hospitalization and death. Use of hospitalization as outcome represents a useful dichotomous outcome that is a proxy for disease severity. However, the decision to admit may be influenced by factors other than the patient’s immediate status, and may be biased toward admission of patients with significant comorbidities, advanced age, or socio-economic considerations. It is conceivable that viral sequence variants might be associated with differential outcomes looking at more granular and direct disease features such as pulmonary radiologic outcomes or specific complications. The use of machine learning produced predictive models with excellent overall performance, particularly for predicting those patients who would not require hospitalization. Although we took significant steps to limit over-learning by models, including testing on two substantial hold-out sets and performing sensitivity analysis with ‘spiked’ datasets, it is possible that these machine learning models might not be generalizable to patients in other geographic regions or in other health systems.

Overall, our results demonstrate substantial sequence variation in SARS-CoV-2 within a single metropolitan area, where the observed sequences represent a substantial fraction of sequence clades that have been observed globally. Viral clade showed a trend toward worse outcomes for patients infected with virus from clade 2 but this result was of borderline statistical significance in our cohort of 190 patients, and potentially confounded by imbalanced distribution of comorbidities between patients infected with the two major clades. Patient demographics and clinical history were strongly predictive of hospitalization, and viral clade information did not substantially improve predictions, suggesting that it contributes minimally to determination of outcome. Further analysis on larger datasets will be needed to determine if viral clade has significant influence on patient outcome.

## Methods

### Subjects, samples, and sequencing

Institutional Review Board (IRB) approval for this study was obtained from the University of Washington, and all research was conducted in compliance with the Declaration of Helsinki. This study was exempted by the IRB from informed consent requirement as a retrospective chart review study. The subset of nasopharyngeal samples collected at University of Washington Medicine (UW Medicine) clinical sites between March 5 and April 8, 2020 that tested positive for SARS-CoV2 by quantitative PCR with Ct < 32 were subjected to whole viral genome sequencing as described previously ^25,26^. In brief extracted RNA from positive specimens was converted to cDNA using random hexamers and sequencing libraries were prepared using Nextera XT or Flex kits (Illumina). Libraries were sequenced on MiSeq, NextSeq or NovaSeq instruments (Illumina) using 1×185, 1×75, or 1×100 runs respectively. Raw reads were processed to generate consensus sequences using a custom bioinformatics pipeline (https://github.com/proychou/hCoV19) that combines de novo assembly and read mapping. Raw reads and consensus sequences were deposited to NCBI SRA and Genbank respectively under BioProject PRJNA610428.

Patients with medical records in the UW Medicine health care system had their health records extracted using manual chart review. Data extracted included demographics (age, sex, ethnicity) as well as co-morbid conditions identified on the documented clinical notes. Finally, clinical outcome measures such as hospitalization and mortality were collected as well. Patients without adequate documentation on comorbidities were coded as unknown (n=4) and not included in calculations for significance.

Demographic and clinical characteristic data were summarized with descriptive statistics including frequencies, percentages and two-sided Pearson chi-square tests with clade type and hospitalization status as outcomes of interest. Fisher’s exact test with mid-p correction and Student’s t-test were used when appropriate in univariate analysis of demographic and clinical characteristics to determine significance. Clopper-Pearson confidence intervals were considered for binomial confidence intervals. In an exploratory analysis, four models of variable selection for multivariate logistic regression were used to determine significance: 1) stepwise Akaike Information Criteria (AIC) ^27^; 2) random forests with area under the receiver operating characteristic curve (AUC) as the parameter of interest ^28^; 3) all univariate significant variables with a p-value <0.1; and 4) all covariates. LASSO regression ^29^ using mean squared error and AUC as the lambda tuner was also performed to understand important predictor variables.

### Missing data

Missing data was handled using multiple imputation by chained equations (MICE) after a sensitivity analysis revealed that the missingness of the data was not completely random (i.e. not MCAR). Recent literature has concluded that the number of imputations should be similar to the percentage of incomplete cases, which in our data is 3.4% ^30,31^. Taken together with the computational expense, a number of 10 imputed datasets was chosen with 20 cycles to reach convergence of the sampling distribution of imputed values ^32^. Finally, all analytic variables with continuous, dichotomous, and categorical data were modelled using predictive mean-matching, Bayesian logistic regression, and Bayesian polytomous regression, respectively.

### Machine learning

For machine learning, several models were built with datasets consisting of combinations of demographics, clinical, clade, and genetic data. Genetic data was included as a vector of sequence variants for each sample. Each dataset was split into train and test sets with 160 and 30 samples respectively. Model selection was run using a nested cross validation (CV) format, with 5 outer folds and leave-one-out CV run on each fold. Model tuning was accomplished using 10000 random sets of hyperparameters for each of 4 model architectures (AdaBoost, Extra Trees, Gradient Boosting, Random Forest). Composite precision-recall curves were produced by merging the 5 outer fold predictions, the area under the precision-recall curves (AUPRC) and area under the receiver operating characteristic (AUROC) were compared, and the parameters that produced the lowest validation bias and highest validation score were chosen. Top 1 performance was similar across the models with the Random Forest slightly outperforming the others. The chosen hyperparameters and model were then used for subsequent testing on the holdout test set.

## Data Availability

All sequence data generated in this study has been uploaded to public databases. All computer programs used in analysis are available on request to the corresponding author.

## Author contributions

KN, CL, AL, PR, ALG, KRJ, and RVG designed these experiments. KN, JZS, PS, PR, JA, AR, PM collected data. KN, CL, AL, PS, PR, ALG, KRR, and RVG analyzed data. KN, CL, AL, PR, ALG, and KRR wrote manuscript.

